# Intrathecal oxytocin for neuropathic pain: A randomized, controlled, cross-over trial

**DOI:** 10.1101/2022.11.16.22282417

**Authors:** James C. Eisenach, Regina S. Curry, Timothy T. Houle

## Abstract

**Objective:** To investigate the effect of intrathecal oxytocin compared to placebo on pain and hypersensitivity in individuals with chronic neuropathic pain.

**Study design:** Randomized, controlled, double-blind cross-over study

**Setting:** Outpatient clinical research unit.

**Subjects:** Individuals between ages of 18 and 70 years with neuropathic pain caudal to the umbilicus for at least 6 months.

**Methods:** Individuals received two blinded intrathecal injections of either oxytocin or saline, separated by at least 7 days, and ongoing neuropathic pain (VAS: visual analog scale) and areas of hypersensitivity were measured at intervals for 4 hours. The primary outcome was VAS pain, analyzed by linear mixed effects model. Secondary outcomes were verbal pain intensity scores at intervals for 7 days and areas of hypersensitivity and elicited pain for 4 hr after injections.

**Results:** The study was stopped early after completion of 5 of 40 subjects planned due to slow recruitment and funding limitations. Pain intensity prior to injection was 4.75 ± 0.99 and modeled pain intensity decreased more after oxytocin than placebo to 1.61 ± 0.87.and 2.49 ± 0.87, respectively (p=0.003). Daily pain scores were lower in the week following injection of oxytocin than saline (2.53 ± 0.89 vs 3.66 ±0.89; p=0.001). Hypersensitivity differed between oxytocin and placebo by small amounts in opposite directions depending on modality tested. There were no study drug related adverse effects.

**Discussion:** Although limited by the small number of subjects studied, oxytocin reduced pain more than placebo in all subjects. Further study of spinal oxytocin in this population is warranted.

## Introduction

Preclinical evidence supports the study of oxytocin to treat neuropathic pain. In rodents, neuropathic injury is accompanied by increased expression of receptors acted upon by oxytocin in both sensory afferents and the spinal cord regions affected by the injury, and recovery from neuropathic hypersensitivity in male and female rodents is temporarily reversed by spinal injection of antagonists of these receptors [1]. Oxytocin uniformly alleviates behavioral evidence of nociception and pain in rodent models of acute and chronic injury-induced pain, with targets identified in both the peripheral and central nervous system [2]. Most pharmacologic studies in rodents employ systemic oxytocin administration at dose orders of magnitude greater than are tested in humans, and human studies, which typically utilize intranasal oxytocin administration, are as likely to show no effect as analgesia [3]. Whether this lack of translation reflects differences in dose or species is unclear.

Given the evidence for spinal sites of action and the rapid removal of oxytocin from the blood after systemic administration, we hypothesized that direct delivery of oxytocin into the central nervous system by intrathecal injection would more likely produce analgesia. A couple of reports indicating analgesia from epidural and intrathecal oxytocin have appeared, although these were performed outside the US without ethical review and oversight by federal drug regulatory agencies for safety, and are therefore not cited here. Following neurotoxicity assessment in animals [4], we obtained Investigational New Drug approval from the US Food and Drug Administration to study intrathecal oxytocin in humans. A Phase 1 dose escalation study of 5-150 μg intrathecal oxytocin did not show side effects [5]. A subsequent randomized, controlled trial comparing intrathecal oxytocin, 100 μg to placebo after hip replacement surgery, while failing to show a reduction in daily worst pain for 2 months postoperatively, showed clinically significant improvements in walking, reduced disability, and more rapid opioid cessation over this time period [6].

The purpose of this study, building upon rationale from basic science and the tolerability of intrathecal oxytocin, is to test whether oxytocin, 100 μg, reduces pain more than placebo using a randomized, controlled, double-blind, cross over design in individuals with neuropathic pain. The primary outcome was visual analog pain intensity score at intervals for 4 hours after injection, with a minimum clinically meaningful difference of 0.9 cm on a 0-10 cm scale, from a pre-injection pain of 4.3. Power analysis supported a sample size of 40 subjects to assess this effect size. Secondary outcomes were areas of hypersensitivity (hyperalgesia and allodynia), if present at intervals for 4 hours after injection and pain scores at intervals for 1 week after intrathecal injection.

## Methods

### Study Population

IRB approval was obtained to recruit up to 44 individuals to obtain 40 evaluable individuals. Equal numbers of men and women were to be recruited, with the following inclusion criteria: age 18 - 70 years, weight < 110 kg, and presence of neuropathic pain for > 6 months, with primary pain area caudal to the umbilicus. Women of child-bearing potential and those < 1 year post-menopausal, had to be practicing highly effective methods of birth control or total abstinence from heterosexual intercourse for a minimum of one full cycle before study drug administration. We excluded individuals with prolonged QTc interval on electrocardiogram (since oxytocin has been reported to prolong QT interval [7], those with hypersensitivity, allergy, or significant reaction to any ingredient of the study drug or lidocaine; those with any disease, diagnosis, or condition (medical or surgical) that, in the opinion of the Principal Investigator, would place the subject at increased risk (active gynecologic disease in which increased tone would be detrimental e.g., uterine fibroids with ongoing bleeding), compromise the subject’s compliance with study procedures, or compromise the quality of the data; women who were pregnant (positive result for serum pregnancy test at screening visit), women who were currently nursing or lactating, or those who had delivered within 2 years; and ongoing pain treatment that included spinal cord stimulators, chronic intrathecal drug therapy, or oral opioid treatment for > 3 months at a current dose of > 100 mg morphine per day or equivalent. Subjects were recruited through pain clinics affiliated with Wake Forest University School of Medicine and by advertising in the community.

### Pre-randomization procedures

On a day at least 3 days before study, patients came to the Clinical Research Unit (CRU) to review the informed consent and to confirm eligibility criteria. A detailed medical history was obtained, an electrocardiogram performed and QTc measured, and blood was drawn from women participants to determine pregnancy status. The research nurse trained participants to reliably rate pain to heating of a 2 cm^2^ Peltier controlled thermode (TSA®, Medoc, Ramat Yishey, Israel) applied to lateral calf. The thermode was heated in a controlled fashion to temperatures between 38° and 51° C for 5 sec using a random staircase method. Areas of hyperalgesia and allodynia were defined using a 225 mN von Frey filament and a cotton wisp, respectively, as previously described [8] and pain intensity in response to the von Frey filament applied in the area of hyperalgesia rated by the subject by marking a line on a 10 cm visual analog pain (VAS) scale with “No Pain” on one end and “Worst Imaginable Pain” on the other. Blood pressure, heart rate, and peripheral oxyhemoglobin saturation were recorded during training.

### Randomization

All individuals received intrathecal saline placebo on one study day and oxytocin on the other study day, with the order randomized using a balanced, block randomization, with block size of 4. A computer-generated randomization was created by an individual not otherwise involved in the study and stored at the Wake Forest Health Sciences Research Pharmacy. Study solutions were prepared and shipped to the Research Pharmacy from Anazao Corporation (Orlando, Florida) under Investigational New Drug approval from the FDA and stored according to manufacturer instructions in the Research Pharmacy until dispensed on the day of study.

### Blinding

The Principal Investigator, research nurse, and subjects were blinded to study drug content. Study drug was provided on each study day in a 3 mL volume in a sterile syringe with label including the name of the study and “Either Saline or Oxytocin, 100 μg”. Blinding could be broken during conduct of the study in the case of a serious adverse event.

### Intrathecal injections

A peripheral intravenous catheter was inserted into a vein in an upper extremity and lactated Ringers solution infused at 1.5 ml/kg/hr for the duration of the study. Patients were positioned either in lateral decubitus or sitting position and the lumbar midline of the back prepped and draped. After obtaining clear, free flow cerebrospinal fluid, a 2 mL sample was obtained and stored for future use under Wake Forest Health Sciences IRB Protocol: 00003383 with the sample stored with a unique identifier. Study solution in a 3 ml volume was then injected over 30 seconds via a #27 or #25 Whitacre spinal needle inserted in a lower lumbar interspace with free flow of cerebrospinal fluid confirmed prior to and after injection. The spinal needle was removed and the subject positioned themselves supine with the head of the bed elevated for the next portion of the study. The two intrathecal injections in this cross-over study were separated by a minimum of 7 days. In the case of inability to successfully place the spinal needle to inject study solution, that study was abandoned.

### Study measures: efficacy

Current pain in the neuropathic area was rated using a 10 cm VAS scale. Areas of allodynia and hyperalgesia were mapped and pain in response to von Frey filament applied in the area of hyperalgesia was rated the VAS scale prior to intrathecal injection, then at 30, 60, 90, 120, 180, and 240 min after injection.

### Safety and Monitoring

#### Study measures: safety

Peripheral oxyhemoglobin saturation, blood pressure, and heart rate were measured non invasively and recorded before and 15, 30, 60, 120, 180, 240, minutes after intrathecal injection. Significant changes were defined as > 30% from baseline in mean arterial pressure or heart rate or oxyhemoglobin saturation by pulse oximetry < 90% or smaller changes if accompanied by side effects. Protocols to treat changes in these monitored parameters were defined in the IRB approved protocol. The electrocardiogram was continuously monitored for the duration of the study, and a tracing printed and the QTc interval calculated at 15 minute intervals for the first hour then hourly until discharge.

A screening neurologic examination was performed at 45, 90, 150, 210, 240 minutes, and 24 hours after intrathecal injection, consisting of deep tendon reflexes, perception of light touch, and extension/withdrawal strength in the arms and legs. In addition subjects were systematically queried at the time of neurologic examination for other symptoms, including sedation, anxiety, nausea, gastrointestinal or bladder discomfort, dizziness, extremity weakness, or any other symptom which was not specifically asked. Any symptom proffered was categorized on a scale of the subject’s choosing.

Oxytocin stimulates both oxytocin and vasopressin receptors, and its action on the latter can result in hyponatremia [9]. Therefore, subjects returned to the CRU 24 hr after each intrathecal injection, for vital sign measurement and a blood draw for serum sodium assay.

A data safety monitoring committee, comprised of professors at Wake Forest University School of Medicine who were not involved in the study, was established prior to starting the study. Blinded data and adverse events were reported to these individuals quarterly. Adverse events were also reported to the IRB and the FDA on an annual basis. Serious adverse events were reported to all of these groups within 24 hr and the study halted until feedback was obtained from each. Data and any adverse events were monitored and reviewed by the principal investigator after treatment of each subject.

#### Discharge from CRU and follow up

After the last study measures were obtained and the intravenous catheter was removed, subjects were discharged to home accompanied by an individual of their choosing where they could be reached by telephone for the remainder of that day, provided that vital signs were within 20% of those on admission and the subject could ambulate without difficulty. They were instructed to call the designated study personnel and were provided with pager and telephone numbers of the research nurse and the investigator should they have any questions or concerns. The research nurse called the subject at 6 hours and 12 hours after intrathecal injection and inquired regarding any problems. Subjects returned to the CRU at 24 hr and were called 2, 3, 4, 5, 7 days after each intrathecal injection, queried for any problems and rated their average pain in the neuropathic area for the past 24 hours using a 0-10 verbal scale. In addition, they were called for these assessments weekly until one month and then at 6 months after the second intrathecal injection.

#### Statistical analysis plan

The intention to treat population was defined as individuals who were randomized and received intrathecal drug on both study days of the trial. The primary outcome measure was pain intensity at rest by VAS on each intrathecal injection study day. Power calculation, based on previously published work by us in this patient population [10, 11], assumed a baseline VAS ongoing pain intensity of 4.3 ± 2.0, a mean reduction in pain 60 min after injection in the study population of 0.9 ± 1.5, and a minimum clinically meaningful group difference of 0.7, resulted in a sample size of 40 with α=0.05 and 1-β=0.80. To account for repeated measures across time and drug conditions, the primary and secondary outcomes were analyzed using a linear mixed-effects model with random intercepts at the level of participant. Each outcome was regressed on fixed-effects for occasion (i.e, order), drug, time, and drug x time interaction. The intra-day models also included pre-drug baseline as a fixed-effect covariate. Main effects are summarized using least square means. P<0.05 was considered significant.

Secondary outcome measures were defined as average verbal pain intensity scores for the previous 24 hr obtained via telephone during the first week after intrathecal injection and areas of hyperalgesia and allodynia and pain elicited by von Frey application in the area of hyperalgesia on each intrathecal injection day. Groups were compared for these outcomes using the linear mixed-models above. Two-tailed hypothesis testing was conducted, with P< 0.05 considered as statistically significant without correction for these multiple secondary outcomes.

Continuous safety variables (vital signs, QTc interval) were compared using repeated measures ANOVA and presence of changes in neurologic testing or symptoms were described. All analyses were conducted using R4.2.1 and OriginPro 2022 (OriginLab, Northampton, MA).

## Results

### Study population

Between June, 2014 and July, 2017, eight subjects were recruited, with the last follow up occurring in January, 2018. Two subjects were not randomized, one because of an abnormal electrocardiogram and the other for lack of spontaneous neuropathic pain at rest. Six subjects were randomized and received the first intrathecal injection. One subject experienced a post-dural puncture headache after the first injection and was followed for adverse event reporting only. As such, there were 5 subjects in the intention to treat population, all of whom completed the study through the 6 month follow up. There were 2 women and 3 men, 2 Black and 3 white, and no Hispanics. Their age, height, and weight (mean ± SD) were 49 ± 10 years, 171 ± 11 cm, and 86 ± 16 kg. The location of neuropathic pain was in the feet or feet and legs in all cases, with diagnosis of peripheral neuropathy due to diabetes in three cases, previous chemotherapy in one case, and previous osteosarcoma in one case. One subject did not have hypersensitivity associated with their neuropathic pain, so the sample size for that outcome was four. Sample size for all other outcomes was five. The time between study drug injections was 7 days in three subjects, 10 days in one, and 22 days in one. The study was stopped early due to cessation of funding.

### Primary analysis

There was a significant effect of study day, drug, and time, but not drug X time interaction on VAS pain in the linear mixed effects model (Table 1). Modeled pain across time and study drug was lower on study day 1 (1.67 ± 0.87 [mean ± SE]) than on study day 2 (2.43 ± 0.87). Modeled pain across time and study day was lower after intrathecal oxytocin (1.61 ± 0.89) than saline (2.49 ± 0.89). The effect of study drug on modeled pain over the 4 hr after intrathecal injection is shown in Figure 1A.

**Table 1.**
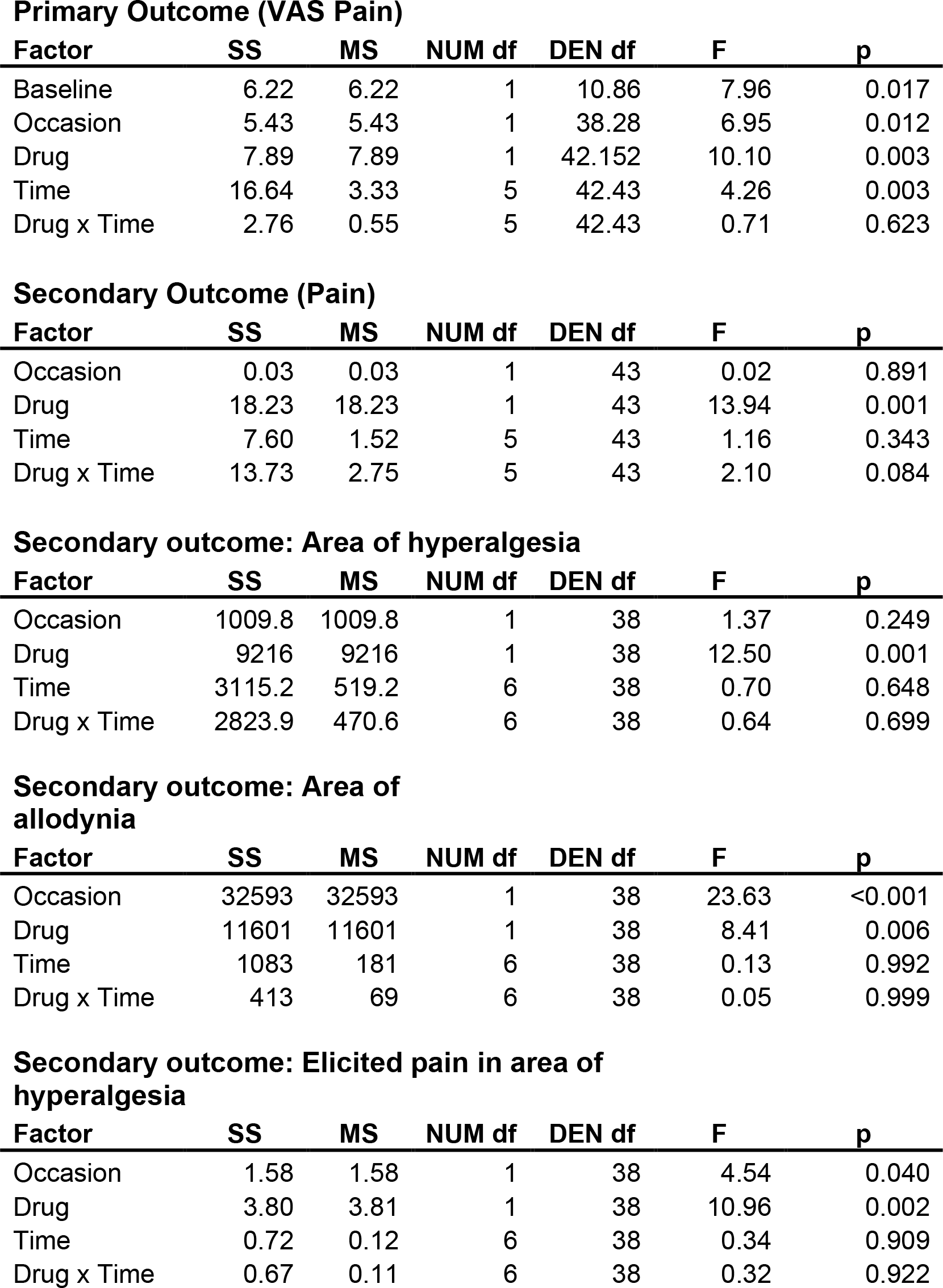
Statistical analysis results. Summary of linear mixed-effects model with random intercepts at the level of participant. SS=sum of squares; MS=mean of squares; NUM df=degrees of freedom in the model; DEN df=number of degrees of freedom associated with model errors.

| **Factor** | **SS** | **MS** | **NUM df** | **DEN df** | **F** | **p** |
| --- | --- | --- | --- | --- | --- | --- |
| Baseline | 6.22 | 6.22 | 1 | 10.86 | 7.96 | 0.017 |
| Occasion | 5.43 | 5.43 | 1 | 38.28 | 6.95 | 0.012 |
| Drug | 7.89 | 7.89 | 1 | 42.152 | 10.10 | 0.003 |
| Time | 16.64 | 3.33 | 5 | 42.43 | 4.26 | 0.003 |
| Drug x Time | 2.76 | 0.55 | 5 | 42.43 | 0.71 | 0.623 |

**Secondary Outcome (Pain)**
| **Factor** | **SS** | **MS** | **NUM df** | **DEN df** | **F** | **p** |
| --- | --- | --- | --- | --- | --- | --- |
| Occasion | 0.03 | 0.03 | 1 | 43 | 0.02 | 0.891 |
| Drug | 18.23 | 18.23 | 1 | 43 | 13.94 | 0.001 |
| Time | 7.60 | 1.52 | 5 | 43 | 1.16 | 0.343 |
| Drug x Time | 13.73 | 2.75 | 5 | 43 | 2.10 | 0.084 |

**Secondary outcome: Area of hyperalgesia**
| **Factor** | **SS** | **MS** | **NUM df** | **DEN df** | **F** | **p** |
| --- | --- | --- | --- | --- | --- | --- |
| Occasion | 1009.8 | 1009.8 | 1 | 38 | 1.37 | 0.249 |
| Drug | 9216 | 9216 | 1 | 38 | 12.50 | 0.001 |
| Time | 3115.2 | 519.2 | 6 | 38 | 0.70 | 0.648 |
| Drug x Time | 2823.9 | 470.6 | 6 | 38 | 0.64 | 0.699 |

**Secondary outcome: Area of allodynia**
| **Factor** | **SS** | **MS** | **NUM df** | **DEN df** | **F** | **p** |
| --- | --- | --- | --- | --- | --- | --- |
| Occasion | 32593 | 32593 | 1 | 38 | 23.63 | <0.001 |
| Drug | 11601 | 11601 | 1 | 38 | 8.41 | 0.006 |
| Time | 1083 | 181 | 6 | 38 | 0.13 | 0.992 |
| Drug x Time | 413 | 69 | 6 | 38 | 0.05 | 0.999 |

**Secondary outcome: Elicited pain in area of hyperalgesia**
| **Factor** | **SS** | **MS** | **NUM df** | **DEN df** | **F** | **p** |
| --- | --- | --- | --- | --- | --- | --- |
| Occasion | 1.58 | 1.58 | 1 | 38 | 4.54 | 0.040 |
| Drug | 3.80 | 3.81 | 1 | 38 | 10.96 | 0.002 |
| Time | 0.72 | 0.12 | 6 | 38 | 0.34 | 0.909 |
| Drug x Time | 0.67 | 0.11 | 6 | 38 | 0.32 | 0.922 |

**Figure 1.**
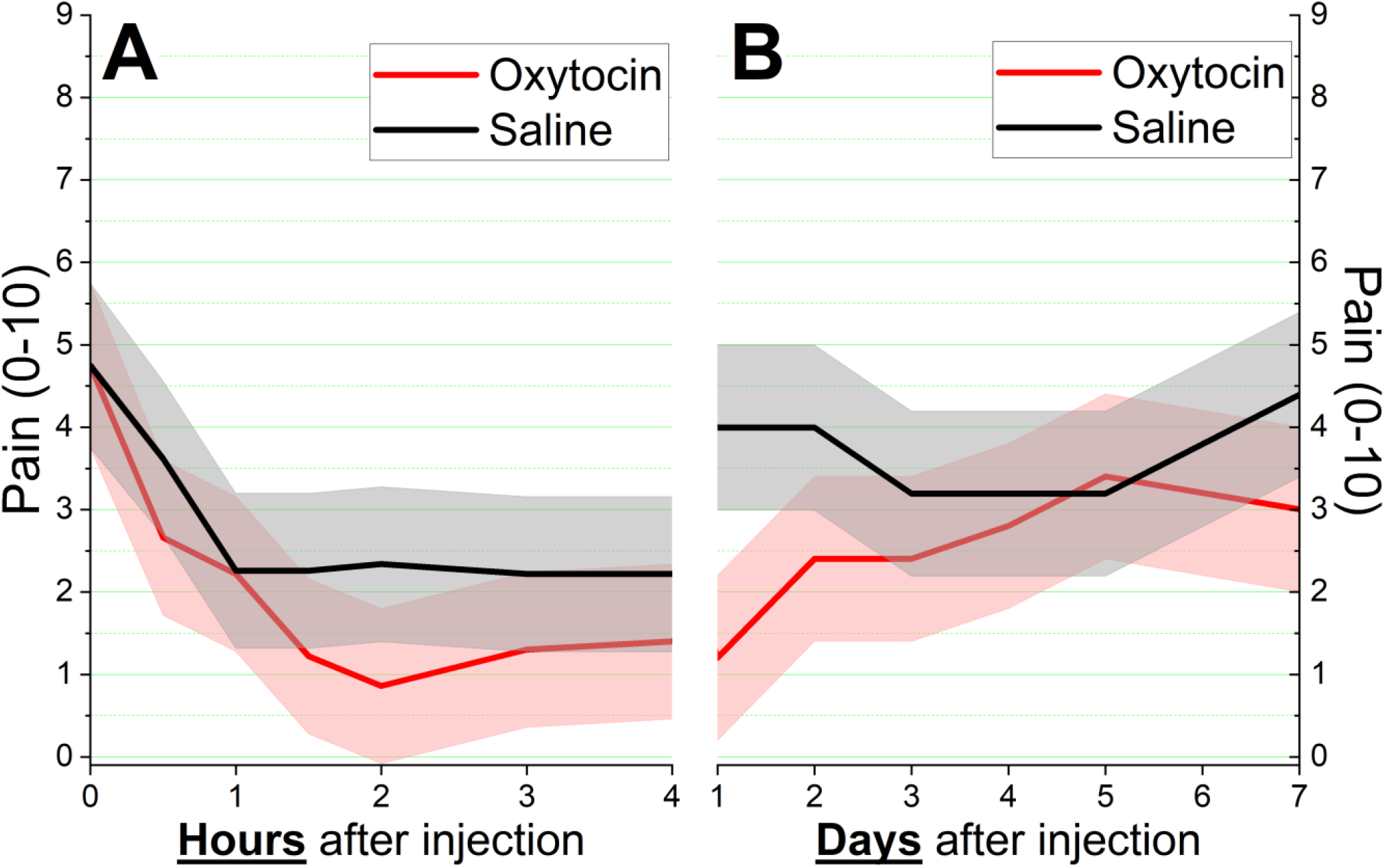
Modeled pain ± SE over **A)** hours and **B)** days after intrathecal injection of oxytocin (red) or saline (black) controlling for order of injection and baseline pain.

### Secondary analyses

#### Neuropathic pain on days after intrathecal injection

In contrast to VAS pain in the 4 hours after injection, average daily pain during the week following injection did not differ by study day or time, but there was a highly significant effect of drug (Table 1). Modeled pain across time was lower after intrathecal oxytocin (2.53 ± 0.89) than saline (3.67 ± 0.89). The effect of study drug on modeled pain over the 7 days after intrathecal injection is shown in Figure 1B. As shown in Figure 2, mean pain scores were lower after oxytocin than saline in every individual regardless of order of injection or duration of time between injections.

**Figure 2.**
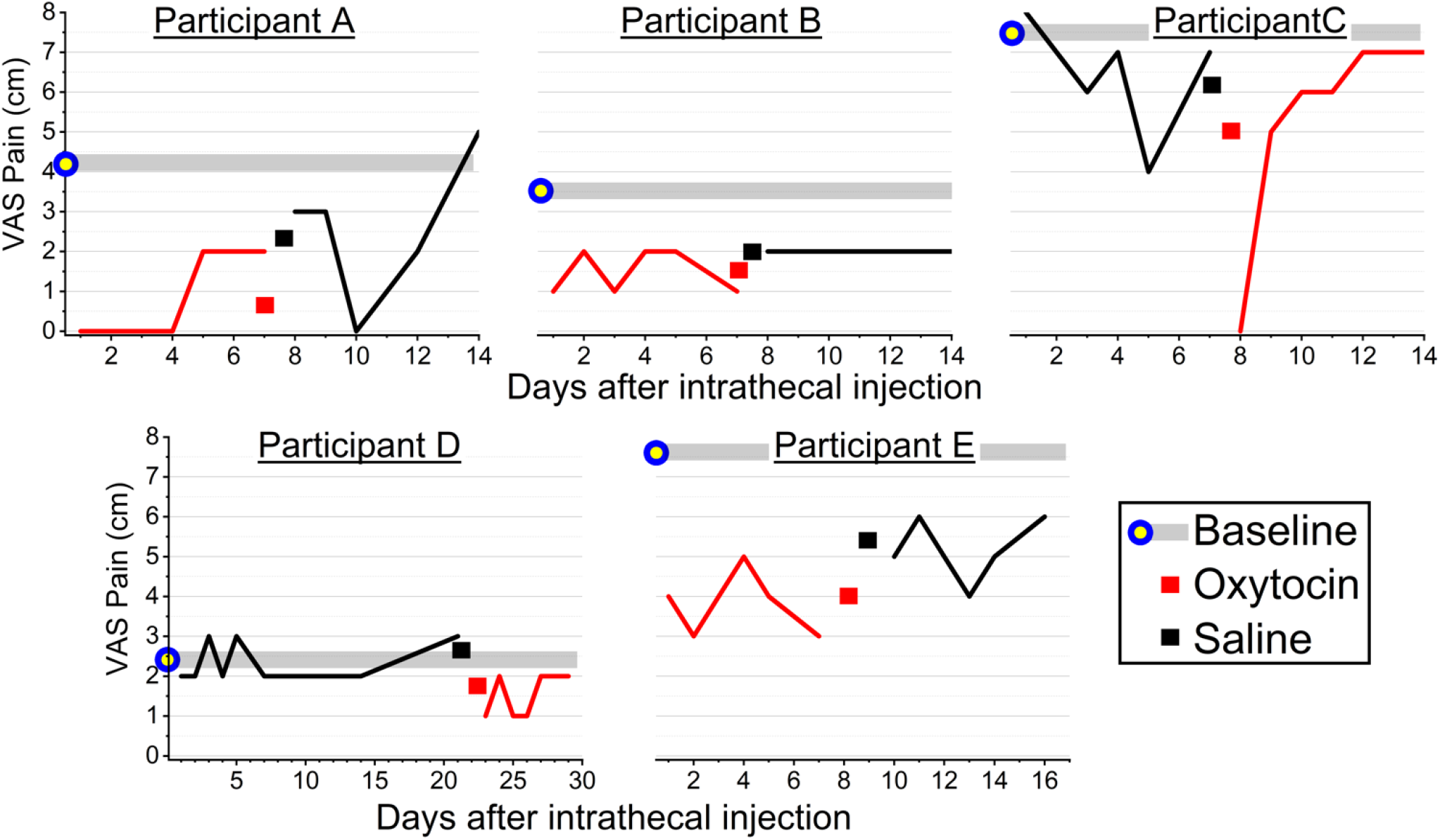
Individual participant average pain in the past 24 hours after discharge from the clinical research unit after receiving oxytocin (red line) or placebo (black line). Note that some subjects received oxytocin first and others received saline first. The red and black squares indicate the mean daily pain scores after oxytocin and saline, respectively, and show that, in each participant, average scores were numerically lower after oxytocin. For descriptive purposes, the visual analog scale pain score at rest prior to the first intrathecal injection is shown in the blue and yellow circle and the horizontal grey band.

#### Areas of hypersensitivity

In the 4 individuals with hypersensitivity accompanying their neuropathic pain, the areas of hyperalgesia and allodynia (average ± SD) prior to oxytocin and saline injection days were similar (182 ± 238 and 197 ± 229 cm^2^, respectively for hyperalgesia and 277 ± 312 and 258 ± 254 cm^2^, respectively for allodynia).

There was a significant effect only for drug on hyperalgesia, but for both drug and study day on allodynia (Table 1). The direction of drug effect differed by test, with modeled area of hyperalgesia being 14% smaller after oxytocin than after saline (181 ± 121 vs 207 ± 121 cm^2^, respectively) in contrast to modeled area of allodynia, which was 11% larger after oxytocin than after saline (272 ± 135 vs 244 ± 135 cm^2^, respectively).

#### Pain within hyperalgesic area

In the 4 individuals with hypersensitivity, pain elicited by von Frey filament application in the hyperalgesic area was similar prior to oxytocin or saline injection (2.8 ± 2.3 and 2.5 ± 1.7 cm^2^, respectively). There was a significant effect of drug and study day on elicited pain in the hyperalgesic area (Table 1), with modeled elicited pain being 18% greater after oxytocin than after saline (2.89 ± 0.89 vs 2.37 ± 0.89, respectively).

#### Safety Monitoring

Neurological sensory, motor, and deep tendon reflexes in the arms and legs were normal before study drug injection and did not change after injection. Mean arterial pressure and heart rate did not differ before oxytocin or saline injection (89 ± 12 vs 91 ± 13 mm Hg and 67 ± 12 vs 67 ± 10 beats per minute, respectively) or after injection (93 ± 11 vs 90 ± 13 mm Hg and 71 ± 9 vs 68 ± 9 beats per minute, respectively; 2-way repeated measures ANOVA p=0.401 for mean arterial pressure and p=0.886 for heart rate, respectively). No subject met criteria for treatment. Oxyhemoglobin saturation averaged 99% before and after injections and there were no measurements <94%. QTc interval did not differ between oxytocin and saline injection days before (414 ± 20 vs 427 ±14 msec, respectively) or after (415 ± 22 vs 425 ± msec, respectively; 2-way repeated measures ANOVA p=0.388). All QTc intervals were within the sex-adjusted normal range. Serum sodium on the day after intrathecal injection did not differ between oxytocin and saline treatments (138 ± 3 vs 138 ± 2 nmol/L, respectively), and all values were within the normal range for our laboratory (134-144 nmol/L).

#### Adverse events

There was one serious adverse event, a pontine infarct which occurred 45 days after the second intrathecal injection accompanied by weakness which partially resolved over the subsequent 4 months. Given the presence of pre-existing medical conditions associated with risk of stroke and the time from study until the event, this was not considered to be due to study drug injection.

One individual had a post dural puncture headache after the first injection (which was saline). The headache was successfully treated with epidural blood patch, this individual did not receive a second injection and was therefore not included in the intent-to-treat population.

One subject reported anxiety before the first intrathecal injection, but no subjects reported anxiety after injections. No subjects reported sedation, gastrointestinal or bladder discomfort, dizziness, or itching before or after injections. One subject reported nausea before and after saline and oxytocin injections, and two subjects reported subjective weakness before and after saline and oxytocin injections. One subject reported a mild headache the day after saline and oxytocin injections which resolved thereafter. Two subjects after saline injection reported mild back ache or stiffness on the day after saline injection which resolved thereafter.

## Discussion

This study demonstrates efficacy of intrathecal oxytocin in five patients with chronic neuropathic pain. Pain intensity scores decreased more by oxytocin than placebo in 100% of the subjects in 4 hours after injection and for the following week, and in both cases the difference exceeded the predefined minimum clinically meaningful difference. The effect size of oxytocin was large (>75% reduction in pain: Figure 1), and there was no evidence of unblinding due to adverse events or subjective sensations. Despite the use of rigorous study design and statistical analysis, however, failure to recruit more than a small fraction of the planned enrollment limits these findings to encouraging further research rather than definitively testing the proposed hypothesis.

Antinociceptive and anti-hypersensitivity efficacy of oxytocin is uniformly observed after systemic and intrathecal injection in rodents [2]. Nearly all studies of oxytocin for analgesia in humans involve systemic administration, most commonly via intranasal spray, with similar numbers of negative and positive results [3]. There is considerable skepticism regarding clinical studies of intranasal oxytocin in neuroscience due to lack of understanding of drug disposition and low scientific rigor of many studies [12]. Thus, translation of pre-clinical findings of intrathecal oxytocin for analgesia to humans has not been adequately examined.

The strong evidence for analgesia in the individuals in this study is in contrast to lack of effect of the same dose of intrathecal oxytocin to alter pain after major orthopedic surgery [6]. There are important distinctions between these two studies which may underlie that difference. Patients after total hip arthroplasty received either intrathecal oxytocin or saline in a randomized, controlled, blinded manner with primary outcome of the modeled worst daily pain during the first 2 postoperative months, and the primary analysis was negative for a difference. Secondary outcomes (ambulation, disability, and time to opioid cessation), however, were all improved in the oxytocin group, suggesting that patients who received oxytocin were more active without the need for more opioids to achieve the same level of worst daily pain. In contrast, pain at rest for a few hours and then <u>average</u> daily pain were assessed in the current study of individuals with neuropathic pain.

Despite reduction in resting pain after intrathecal oxytocin, we observed mixed effects on hypersensitivity associated with neuropathic pain, with area of hyperalgesia slightly reduced but area of allodynia and elicited pain in the hyperalgesic area slightly increased with oxytocin compared to placebo. This is noteworthy from a preclinical translation standpoint, since most rodent studies of oxytocin, including those we have performed, rely primarily on hypersensitivity to mechanical stimuli as a correlate of pain and for assessment of analgesia.

There were no side effects observed ascribed to study drug in these patients with neuropathic pain as there were no side effects from this dose of intrathecal oxytocin in patients undergoing total hip arthroplasty [6] and of larger oxytocin doses in healthy volunteers [5]. Yet, the number of people who have received intrathecal oxytocin remains very small and this precludes clinical application without further systematic safety assessments.

There are several limitations to this study. A very small sample size is associated with higher likelihood of false positive findings and over-estimation of effect sizes [12]. Only resting pain was assessed on the study day, and level of activity or disability because of neuropathic pain was not assessed after discharge from the clinical research unit. Only people with peripheral neuropathy in the lower extremities from diabetes or other causes were studied, limiting generalizability.

## Conclusions

In summary, intrathecal oxytocin was well tolerated by the 5 individuals studied and, in each individual, intrathecal oxytocin reduced pain by a clinically meaningful amount but had mixed effects on hypersensitivity in the hours and days following injection compared to saline placebo. These results encourage further study of oxytocin in treatment of neuropathic pain.

## Data Availability

All data produced in the present study are available upon reasonable request to the authors

